# Machine Learning in Clinical Psychology and Psychotherapy Education: A Survey of Postgraduate Students at a Swiss University

**DOI:** 10.1101/2020.11.15.20231308

**Authors:** Charlotte Blease, Anna Kharko, Marco Annoni, Jens Gaab, Cosima Locher

## Abstract

**Background:** There is increasing use of for machine learning-enabled tools (e.g., psychotherapy apps) in mental health care.

**Objective:** This study aimed to explore postgraduate clinical psychology and psychotherapy students’ familiarity and formal exposure to topics related to artificial intelligence and machine learning (AI/ML) during their studies.

**Methods:** In April-June 2020, we conducted a mixed-methods web-based survey using a convenience sample of 120 clinical psychology and psychotherapy enrolled in a two-year Masters’ program students at a Swiss university.

**Results:** In total 37 students responded (response rate: 37/120, 31%). Among the respondents, 73% (n=27) intended to enter a mental health profession. Among the students 97% reported that they had heard of the term ‘machine learning,’ and 78% reported that they were familiar with the concept of ‘big data analytics’. Students estimated 18.61/3600 hours, or 0.52% of their program would be spent on AI/ML education. Around half (46%) reported that they intended to learn about AI/ML as it pertained to mental health care. On 5-point Likert scale, students moderately agreed (median=4) that AI/M should be part of clinical psychology/psychotherapy education.

**Conclusions:** Education programs in clinical psychology/psychotherapy may lag developments in AI/ML-enabled tools in mental healthcare. This survey of postgraduate clinical psychology and psychotherapy students raises questions about how curricula could be enhanced to better prepare clinical psychology/psychotherapy trainees to engage in constructive debate about ethical and evidence-based issues pertaining to AI/ML tools, and in guiding patients on the use of online mental health services and apps.

## Introduction

### Background

Digital services based on artificial intelligence and machine learning (AI/ML) are increasingly used in mental health care via the use of apps. Health apps encompass a range of proposed uses, including the monitoring and tracking of symptoms, as well as direct-to-consumer interventions designed to support, complement, or replace, psychotherapy (Firth and Torous, 2015; Lui et al., 2017). Psychotherapy apps have been designed to include various techniques including cognitive behavioral therapy, acceptance commitment therapy, and eclectic therapy. The recent coronavirus crisis has further accelerated the shift toward a model in which therapeutic relationships are increasingly mediated by on-line platforms and digital services.

Considering these digital advances, educating future clinicians, including psychologists and psychotherapists, will be important to ensure optimal, safe use of AI/ML enabled tools and innovations. So far, a growing number of investigations have explored the views of clinicians including primary care physicians on the impact of AI/ML tools on their job (Boeldt et al., 2015; Blease et al., 2018, 2019a). These studies, albeit limited, suggest that mental health clinicians expect AI/ML to influence or change their professional roles in the future. For example, in 2020, an international survey of 791 psychiatrists reported that 75% (n= 593) believed that AI/ML enabled tools would, at some point, be able to fully replace psychiatrists in documenting and updating clinical records (Doraiswamy et al., 2020). In the same survey, 54% (n=427) of psychiatrists believed that AI/ML tools will be able to fully replace humans in synthesizing information to make diagnoses (Doraiswamy et al., 2020). In qualitative research, psychiatrists express divergent opinions on the benefits and harms of AI/ML in treating mental health patients with comments demonstrating scarce reflection of ethical and regulatory considerations for patient care (Blease et al., 2019b). Similarly, in a recent survey of psychiatrists in France (n=515) (Bourla et al., 2018), respondents expressed “moderate acceptability” of disruptive technologies, such as wrist bands for monitoring symptoms, but concluded that this likely reflected lack of extensive knowledge about these technologies. However, we are not aware of any surveys that have explored the opinions, openness, and familiarity of clinical psychology and psychotherapy students on the impact of AI/ML on their future job. In addition, only limited research has explored the awareness, opinions, and formal education of medical students on these topics (Dos Santos et al., 2018).

### Objectives

We aimed to expand existing research by exploring whether clinical psychology and psychotherapy students believed their career choice would be impacted by AI/ML, the benefits and harms of any such impact, and their level of formal training on these topics. Our goal was to explore whether more education may be required so that trainee clinical psychologists/psychotherapists might harness these tools in the future.

## Methods

### Study population

The single-center study was based at the Faculty of Psychology, University of Basel, Switzerland. The online survey was conducted April to June 2020 with clinical psychology and psychotherapy students (see Supplementary File 1). Students were first- and second-year post-graduate students enrolled on a two-year Masters’ degree program in clinical psychology and psychotherapy. =

Respondents enrolled in the Masters’ program were invited via email to participate in the study. Three further reminder emails were sent, one to two weeks apart. Participation was voluntary and students were advised that the survey was not a test, that their responses would be pseudonymized, and that no sensitive information would be collected. There was no selection or exclusion in recruitment, and no reimbursement or compensation. Ethical approval for the study was granted by the Faculty of Psychology, University of Basel. The survey was administered in English, as students enrolled on the clinical psychology/psychotherapy Masters’ program at the University of Basel are expected to be fluent English-speakers.

### Survey instrument

The online survey [see Supplementary File 1] was designed with the online software Jisc (https://www.jisc.ac.uk/). Questions were included after consultations with psychotherapists and pre-tested with students from outside the university to ensure face validity and feasibility. The survey opened with a brief statement: “We are inviting you, as psychology students to give your opinions about technology and the future of mental health care.” We also made it clear that the survey was aimed at assessing their personal opinions. We stated that we did not assume that participants have any expertise about AI/ML. In the first section, respondents were asked to provide demographic information. Participants were also requested to state whether they intend to enter a mental health profession or not. The second section consisted of questions encompassing open comment questions on the future of psychotherapy (i.e., respondents had to briefly describe way(s) in which future technology might change psychotherapists’ job in the next 25 years), as well as potential benefits and risks of future technology in treating patients with mental health problems. A second publication will report the qualitative research findings from open comment responses. The third section of the survey was intended to gauge participants’ familiarity with artificial intelligence and machine learning. First, participants had to answer whether they are familiar with ‘machine learning’ and ‘big data analytics’ and whether they read academic journal articles relating to these topics (‘no’, ‘yes’ answers). Participants were requested to estimate the amount time (a) already spent and (b) anticipated on these topics in their program of study. Finally, respondents were also asked to rate the importance of AI/ML for clinical psychology/psychotherapy education.

### Data management and analysis

We used descriptive statistics to examine students’ characteristics and opinions about the impact of AI/ML on the future of psychotherapy. The quantitative data was analyzed to extract summary statistics and 95% confidence intervals. Spearman’s correlation coefficient was calculated for key variables describing students’ experiences and attitudes towards including education about AI/ML in a clinical psychology program.

## Results

### Respondent characteristics

Descriptive statistics and analysis were carried out using JASP (0.9.2). Table 1 provides a summary of demographic characteristics. The final respondent sample comprised 37 students (response rate: 37/120, 31%). There was a homogeneous distribution of students in terms of their current study semester.

**Table 1.** Sample Characteristics (n = 37)

| | $\mu$ or n | (SD) or % |
| --- | --- | --- |
| Gender (female) | 30 | 81% |
| Age (n years) * | 26.65 | (5.21) |
| Year |  |  |
| 1<sup>st</sup> | 24 | 65% |
| 2<sup>nd</sup> | 13 | 35% |
| n intend to enter a mental health profession |  |  |
| Yes | 27 | 73% |
| No | 3 | 8% |
| Unsure | 7 | 19% |
| Of those who said 'Yes' (n = 27), n intend to enter... |  |  |
| Clinical Psychology/Psychotherapy | 23 | 85% |
| Counselling/Coaching | 2 | 7% |
| Social Work | 1 | 4% |
| Other: Neuropsychology | 1 | 4% |

$\mu$ - average value, n – count, SD – standard deviation, % - percentage.
\* Items, for which $\mu$ and SD were calculated.

### Participants’ opinions about, and familiarity, with AI/ML

Results of the quantitative section are shown in Table 2. The vast majority (97%, n=36) had heard of ‘machine learning’ and were familiar with ‘big data analytics’ (78%, n=29). Respondents reported an average (mean) of 6.18 hours, so far, of AI/ML in their degree. They anticipated, on average (mean), a further 12.43 hours of AI/ML education in their Masters’ degree program. While almost half (46%) of surveyed participants reported their intention to learn more about AI/ML as it pertains to mental healthcare, the remaining respondents were either unsure (43%) or responded that they had no intention of doing so (11%).

**Table 2.** AI/ML Education Experience & Interest

|  | m or n | (SD) or % | Range |
| --- | --- | --- | --- |
| *n* have heard of machine learning | 36 | 97% | - |
| *n* are familiar with big data analytics | 29 | 78% | - |
| *n* have read AI/ML mental health journal articles | 23 | 62% | - |
| AI/ML education during the degree (n hours) * |  |  |  |
| *So far* | 6.18 | (16.63) | 0 – 100 |
| *Predicted* | 12.43 | (16.39) | 0 – 60 |
| Intend to learn about AI/ML as it pertains to mental health care |  |  |  |
| *Yes* | 17 | 46% | - |
| *No* | 4 | 11% | - |
| *Unsure* | 16 | 43% | - |
| Discussion about AI/ML should be part of clinical psychology education. * ** | 4 (Moderately agree) | (1.48) |  |

m – mean, n – count, *SD* – standard deviation, % - percentage.
\* Items, for which m and *SD* were calculated.
\*\* Answer was a rating on a 5-point agreement Likert scale, where 1) *Strongly disagree*, 2) *Moderately disagree*, 3) *Neutral*, 4) *Moderately agree* and 5) *Strongly agree*.

Students who intended to learn more about the application of AI/ML in mental health reported more hours of relevant education (m = 9.24) than those who were uncertain (m = 4.44) (see Figure 1). Furthermore, students who intended to learn more stated that they will have more hours of such education in the future (m = 20.88) compared with those who were unsure (m = 6.53) (see Figure 2). Overall, however, students moderately agreed that discussions about artificial intelligence/machine learning should be part of clinical psychology/psychotherapy education.

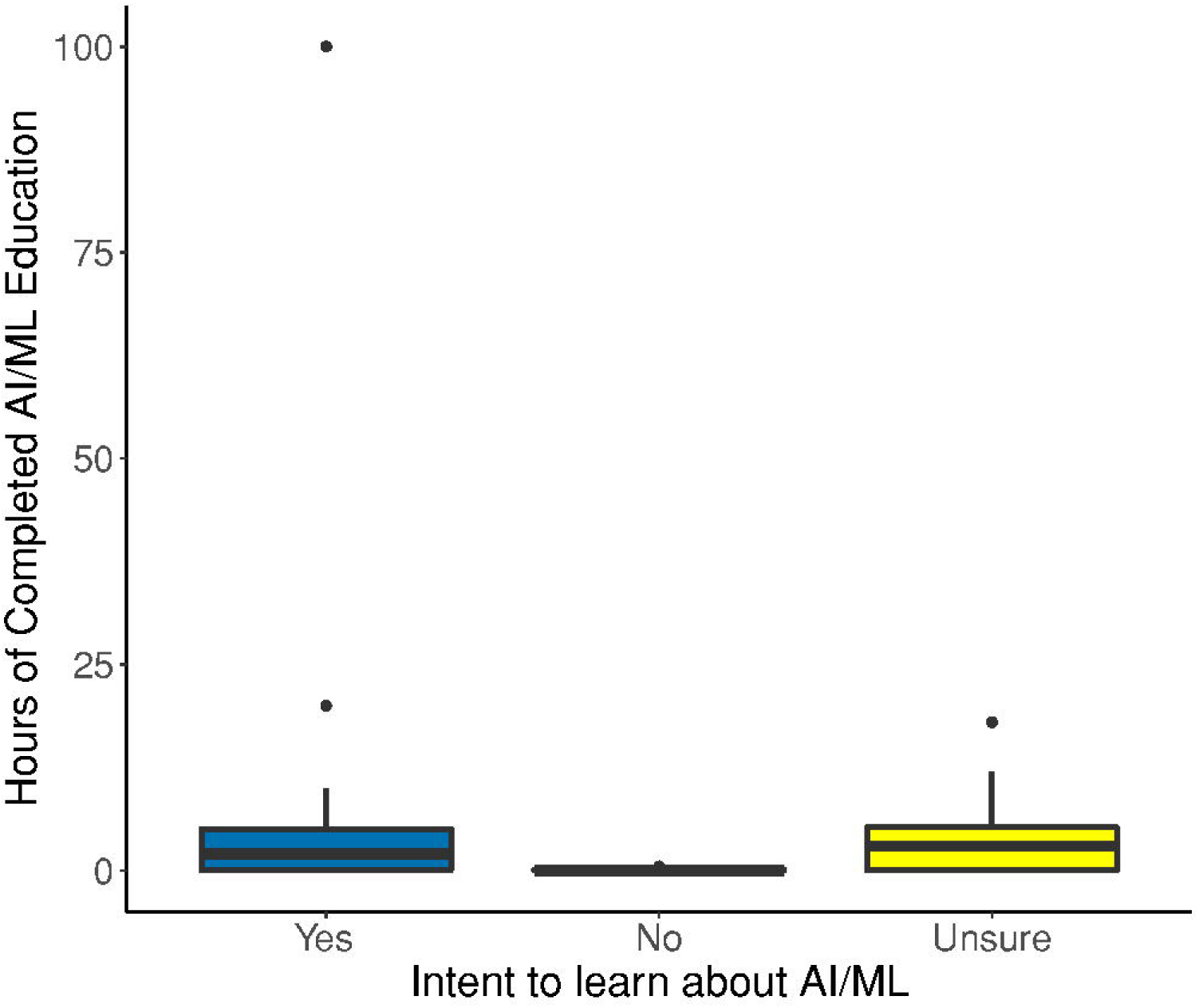

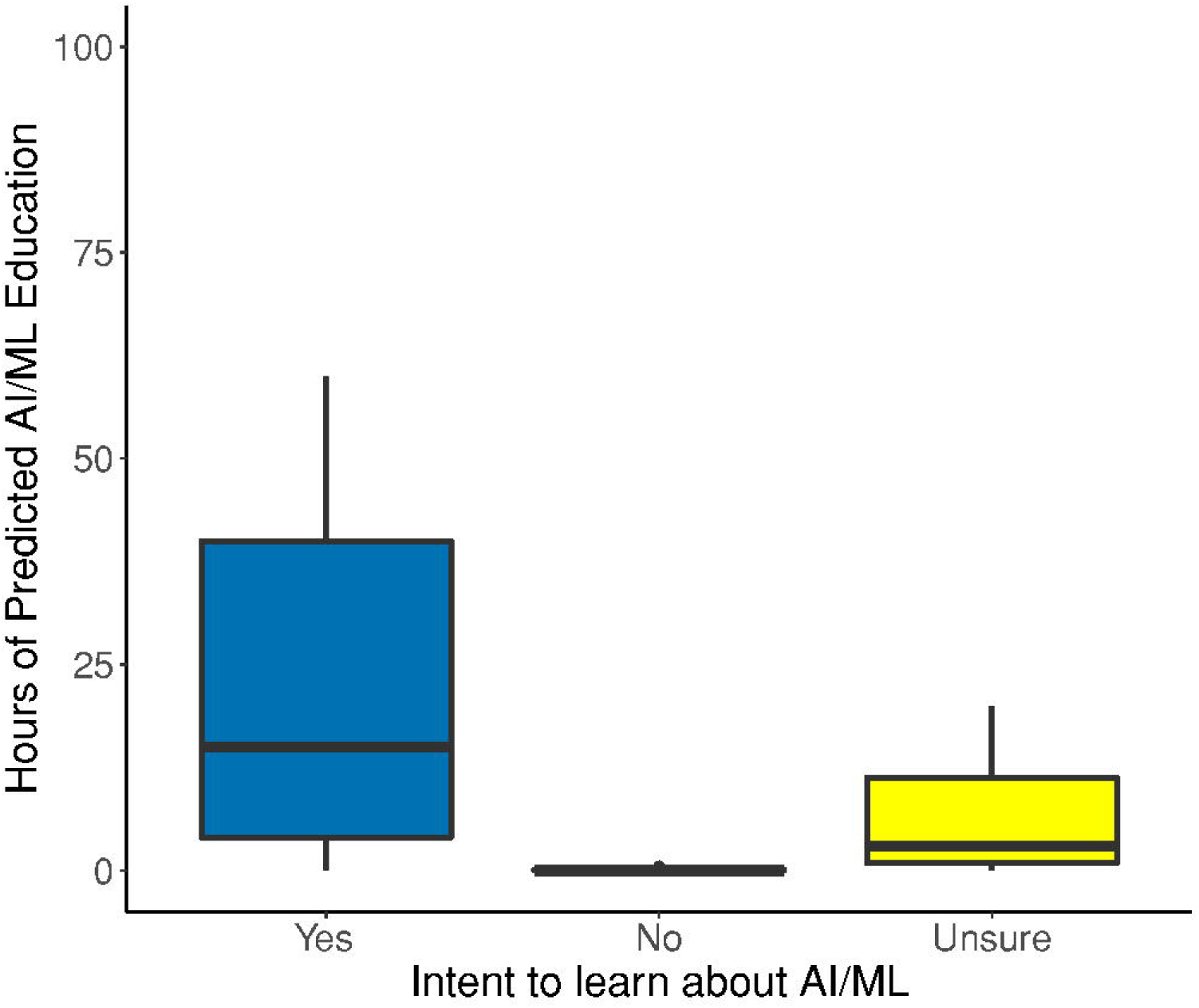

### Correlates of opinions

The only significant positive relationship between respondents’ attitudes about the inclusion of AI/ML in education, and hours spent receiving relevant education. Students who reported receiving more hours of AI/ML education gave a higher rating on the 5-point Likert scale (r = .34, p = .038).

## Discussion

### Summary of major findings

Most postgraduate students in our sample intended to enter a mental health profession, and most had some familiarity with the terms ‘machine learning’ and ‘big data’. Around two thirds of respondents also reported reading a journal article on AI/ML. Around half (46%) the respondents reported their intention to learn more about AI/ML; remaining respondents were unsure, and around one in ten reported no intention of doing so. Respondents also reported receiving an average of 6.18 hours learning, so far, on the topic of AI/ML in their course and expected an average of a further 12.43 hours of teaching on the topic in their degree program. Combining both reported and anticipated time on AI/ML education, this amounts to a perceived total of 18.61/3600 hours, or 0.52% of their total degree.

The opinions and experiences of trainee clinicians have been missing from the debate about the impact of AI/ML on clinical psychology and psychotherapy. This exploratory survey indicates that clinical psychology and psychotherapy students express some awareness of AI/ML. However, students expressed mixed levels of interest to learn more. This may reflect lack of knowledge about how AI/ML is already encroaching on mental health care. In addition, it is possible that students’ current familiarity may be driven less by formal training than by outside sources, including the media.

Reflecting on these findings, the important question arises about whether teaching bodies should be adapted, not only for students but also for educators. In a recent survey, leading healthcare informaticians forecast that by 2029, AI/ML will incur workplace changes in primary care, with the need for increased training requirements in these fields (Blease et al., 2020). As the digital app economy continues to boom there is considerable promise, but also the potential for harm. To date, it is estimated that there are more than 10,000 health apps available for download, yet most have never been subject to robust standards of evidence-based medicine (Lui et al., 2017; Torous and Roberts, 2017; Baumel et al., 2020). Connected to regulatory considerations in the use of psychotherapy apps are ethical issues (Martinez-Martin and Kreitmair, 2018). Loss of privacy, misuse of sensitive healthcare information remains a risk, with known cases of mobile technologies selling patient data to third parties (O’neil, 2016; Cohen and Mello, 2018; Zuboff, 2019). Hence, while there is considerable scope for mobile health innovations in improving patient care (Porras-Segovia et al., 2020; Tseng et al., 2020), there is also a pressing need to formulate clear recommendations for these apps among patients and clinicians. Mental health patients remain among the most of vulnerable patient populations and are especially at risk of privacy violations via the exploitation of their data. This survey raises questions about the preparedness of clinical psychology/psychotherapy students to fully engage in pressing debates about ethical and evidence-based issues pertaining to AI/ML tools, and in guiding patients on the use of psychotherapy and other mental health apps.

### Strengths and limitations

To our knowledge, this is the first survey that aims to investigate the exposure, and opinions of, clinical psychology/psychotherapy students to AI/ML. Although the response rate was moderate, the sample size was small. It is also unknown if response biases affected findings, and whether the decision to complete the survey was influenced by students’ prior knowledge or awareness of the topic of AI/ML. The response rate, and the convenience sample, also raise questions about representativeness.

Some items on the survey could be challenged on the grounds of vagueness. For example, ‘familiarity with big data analytics’ might, justifiably, be considered semantically opaque. While we acknowledge that this survey item is coarse-grained, this preliminary study set out to explore general student awareness, level of personal inquiry, and formal educational exposure to the topic of AI/ML. We recommend that interviews, or focus groups would provide finer-grained analysis of student awareness and opinions of AI/ML. Further, we suggest that future research might usefully explore the views of specific groups of students (for example, only those who aim to work as psychotherapists), and on the views of clinical psychology/psychotherapy, and other mental health educators. In addition, it would be useful to evaluate course curricula across tertiary level colleges and universities to obtain a more objective assessment of topics and level of education about AI/ML in clinical psychology/psychotherapy training.

Finally, students responded to the survey during April to June 2020, during the coronavirus pandemic, and it is not known how, or whether, contextual conditions influenced their responses to the survey. With the recent the uptick in telemedicine, and considerable debate about digital health during the pandemic, it is conceivable that participants’ answers may have been influenced by both global and local conditions.

## Conclusions

Clinical psychologists/psychotherapists entering the job-market will face new challenges posed by the emergence of new e-health tools based on artificial intelligence, machine learning and big data analytics. Although the majority of students in our survey had heard of ‘machine learning’ and read about AI/ML in journal articles, only half of respondents planned to learn more about AI/ML as they pertain to mental health care. Importantly, most students agreed that discussions about AI/ML should be part of clinical psychology/psychotherapy education. Yet they estimated only 0.52% of their total degree (18.61/3600 hours) will be dedicated to these topics. These results seem to contrast with current trends. Clinical psychologists/psychotherapists – as well as patients/clients – can already access thousands of digital tools, online services and mobile apps based on AI/ML that have been specifically designed to integrate or substitute traditional mental healthcare services or consultations. The impact of these technologies on mental healthcare is set to rise as new and more advanced AI/ML tools and services are released.

We suggest that clinical psychology/psychotherapy curricula should embrace these new challenges in educating the clinicians of tomorrow. Courses might be usefully designed to train clinical psychologists and psychotherapists on how to guide and assist patents in being ‘digitally savvy’ – and in making informed choices about available AI/ML tools and services. AI/ML technologies, in fact, raise significant ethical concerns related to their evidence-based effectiveness and safety as well as about other delicate ethical and regulatory issues related to privacy, equality, and discrimination. Course curricula should encompass instruction about these issues so that practitioners can feel empowered to keep abreast of new technological advances including what these developments mean for their profession and their patients.

## Data Availability

The raw data supporting the conclusions of this article will be made available by the authors, without undue reservation.

## Acknowledgements

We would like to express our gratitude to the students who participated in this survey. The authors also thank Dr John Torous for feedback on an earlier draft of this manuscript.

