## Supplementary File 1 for "Machine Learning in Clinical Psychology and Psychotherapy Education: A Survey of Postgraduate Students at a Swiss University"

### MACHINE LEARNING AND THE FUTURE OF MENTAL HEALTH CARE

#### Welcome

**Dear Student,**

**Researchers at the Division of Clinical Psychology and Psychotherapy, University of Basel invite you to take part in a survey. We are inviting you, as psychology students to give your opinions about technology and the future of mental health care. This research will help to inform clinical psychology educationalists and policy-makers.**

The online survey should take around 10 (and no more than 15) minutes to complete. We will not collect any identifying information from you, and your responses will be pseudonymized. Your response will be collated with those of other respondents in aggregated, pseudonymized form. **This survey is not a test; we are interested in your opinions as clinical psychology and psychotherapy students.** We refer you to the Information sheet and consent form for more information.

If you decide to participate, we appreciate your time and contribution to our research.

Thank you.

Dr Cosima Locher, University of Basel

#### OUR TEAM

| | | | |
|---|---|---|---|
| 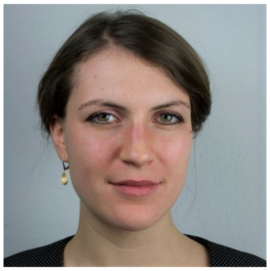 | 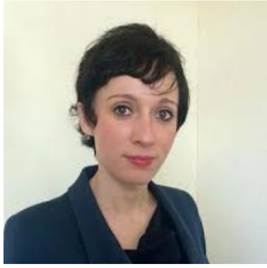 | 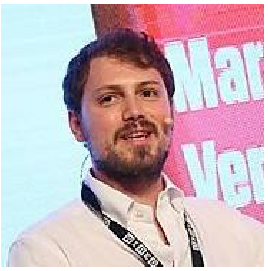 | 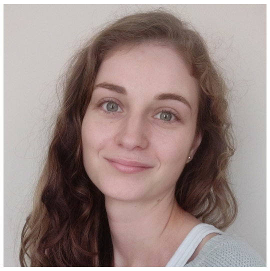 |
| **Dr. Cosima Locher** | **Dr. Charlotte Blease** | **Dr. Marco Annoni** | **Anna Kharko** |
| *Research Fellow* | *Research Fellow* | *Research Fellow* | *PhD Student* |
| Division of Clinical Psychology & Psychotherapy | Beth Israel Deaconess Medical Center | Institute of Biomedical Technologies | School of Psychology |
| University of Basel | Harvard Medical School | National Research Council of Italy | University of Plymouth, UK |

If you have any queries about this research and its results, you can contact Dr. Marco Annoni.

### INFORMATION SHEET

#### Research Objective

Thank you for considering participating in this research project. The purpose of this page is to explain to you what the work is about and what your participation would involve, so as to enable you to make an informed choice.

The purpose of this study is to investigate the opinions of clinical psychology and psychotherapy students regarding the potential impact of machine learning/artificial intelligence on the future of mental health care and psychotherapy. We do not expect any knowledge of technologies: instead we are interested in your views as students'.

#### Study Procedure

Should you choose to participate, you will be asked some non-identifying demographic information, followed by open-ended comment boxes asking you to express your opinions on how mental health care and psychotherapy might be impacted by technology. The survey also asks what exposure you have had to information about artificial intelligence in your degree program.

#### Duration & Compensation

The survey will take around 10-15 minutes to complete. You will receive '1 Unterschrift' for the participation in this study.

#### Risks & Benefits

By participating in this study you have the possibility to gain insight into psychological research. Furthermore, your responses will help us to better understand the opinions of future clinical psychology and psychotherapy professions on artificial intelligence, which in turn may help to inform clinical psychology curricula. While there are no direct benefits to you, participating in this survey may help to stimulate you to think about the role of technology in healthcare. This study entails no risks that would exceed the risks ordinarily encountered in daily life. We do not anticipate any negative outcomes from participating in this study.

#### Voluntary Participation

Participation in this study is completely voluntary. There is no obligation to participate, and should you choose to do so you can refuse to answer specific questions, or decide to withdraw from the study. We will not collect any personal or sensitive information therefore the survey will be fully anonymous. The survey is not a test, and your decision to participate will not affect your grades. Ticking the box below will indicate consent to participate.

You maintain the right to withdraw from the study at any stage up to the point of data submission. At this point your data will be collated with that of other participants and can no longer be retracted.

The anonymous data will be stored securely for up to ten years on the University of Basel Server in secured form. The information you provide may contribute to research publications and/or conference presentations which may be publicly available. However, your contributions will be fully anonymous.

The study has obtained ethical approval from the Research Ethics Committee at the University of Basel, Faculty of Psychology.

If you have any queries about this research and its results, you can contact Dr Cosima Locher. If you agree to take part in this study, please complete the consent form below.

Under these conditions, do you consent to particapte in this study? ✱ *Required*

- ○ Yes, I agree to take part.
- ○ No, I don't agree to take part.

### SECTION A

We store your data securely by using a Participant ID. To create your unique 4-letter PID, please use the instructions below. ✱ *Required*

More info

ABCD

Gender ✱ *Required*

- Female
- Male
- Other
- No information

If you selected Other, please specify:

Year of Birth ✱ *Required*

Current semester of your Masters degree. ✱ *Required*

- 1st
- 2nd
- 3rd
- 4th

What was your undergraduate degree subject(s)? *Please enter.* ✱ *Required*

Do you intend to enter a mental health profession? ✱ *Required*

- ○ Yes
- ○ No
- ○ Unsure

Please select one area only.

- ○ Clinical Psychology/Psychotherapy
- ○ Social Work
- ○ Counselling/Coach
- ○ Other

If you selected Other, please specify:

### SECTION B

The questions in this section are on your opinions about the impact of machine learning/artificial intelligence on the future of mental health care. We do not assume you have any expertise about machine learning or artificial intelligence in health care.

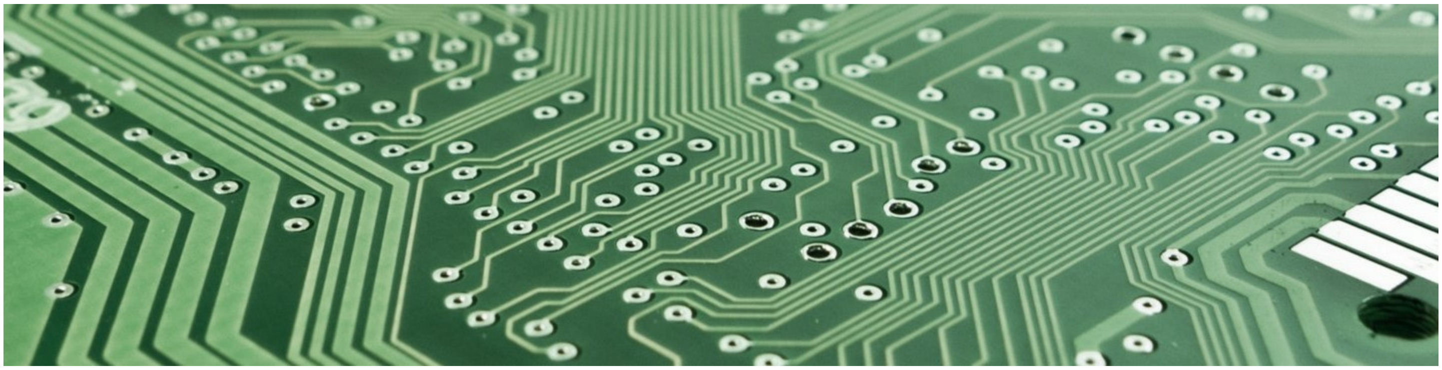

In the next 25 years, please briefly describe the way(s) you believe artificial intelligence/machine learning might change the care of patients with mental health conditions.

In the next 25 years, please briefly describe the way(s) you believe artificial intelligence/machine learning might change the job of clinical psychologists and psychotherapists.

Please provide any brief comments you may have about the potential benefits of artificial intelligence/machine learning to the care of patients with mental health conditions.

Please provide any brief comments you may have about the potential risks of artificial intelligence/machine learning to the care of patients with mental health conditions.

### SECTION C

The questions in this section ask about your familiarity with artificial intelligence/machine learning.

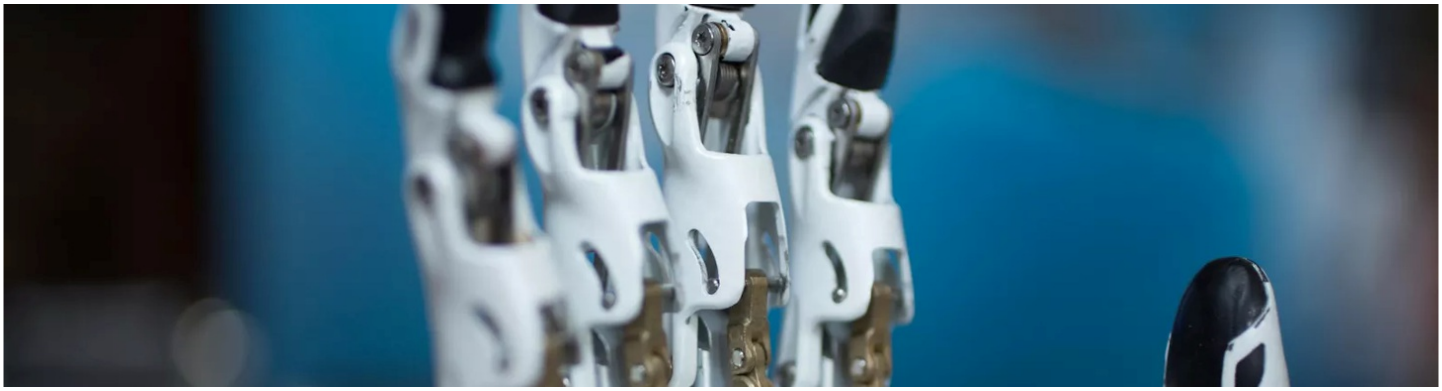

Have you heard of machine learning? ✱ *Required*

- Yes
- No

Are you familiar with big data analytics? ✱ *Required*

- Yes
- No

Have you read any academic journal articles about artificial intelligence/machine learning in mental health care either during your studies or by yourself? ✱ *Required*

- Yes
- No

Please estimate how many hours your instructors/lecturers have spent discussing artificial intelligence/machine learning during your Masters degree so far. ✱ *Required*

Please estimate how many hours your instructors/lecturers will spend discussing artificial intelligence/machine learning during the course of obtaining your Masters degree. ✱ *Required*

Do you plan to learn about artificial intelligence/machine learning as they pertain to mental health care? ✱ *Required*

- Yes
- No
- Maybe

Discussion about artificial intelligence/machine learning should be part of clinical psychology/psychotherapy education. ✱ *Required*

- Strongly disagree
- Moderately disagree
- Somewhat disagree
- Moderately agree
- Strongly agree

#### Questions & Comments

##### Feedback

Do you have any questions for the research team or comments about the topic of the survey? If you do, please share them below. **Otherwise, please click 'Finish'.**

### Final page

#### THANK YOU

Thank you from the research team!

If you have any questions, comments or concerns, please

---

The photographic materials used in this survey are sourced from pexels.com

---
